# Quantifying the effect of isolation and negative certification on COVID-19 transmission

**DOI:** 10.1101/2022.02.20.22270449

**Authors:** Jun-ichi Takeshita, Michio Murakami, Masashi Kamo, Wataru Naito, Tetsuo Yasutaka, Seiya Imoto

**Affiliations:** Research Institute of Science for Safety and Sustainability, National Institute of Advanced Industrial Science and Technology (AIST), Tsukuba, Japan; Center for Infectious Disease Education and Research, Osaka University, Osaka, Japan; Research Institute for Geo-Resources and Environment, Geological Survey of Japan, National Institute of Advanced Industrial Science and Technology (AIST), Tsukuba, Japan; Division of Health Medical Intelligence, Human Genome Center, The Institute of Medical Science, The University of Tokyo, Tokyo, Japan

**Keywords:** SEIR model, Infection probability, Conditional probability, PCR test, Antigen test, Isolation period, Negative test certification, COVID-19

## Abstract

Isolation of close contact people and negative test certification are used to manage the spread of new coronavirus infections worldwide. These effectively prevent the spread of infection in advance, but they can lead to a decline in socio-economic activity. Thus, the present study quantified the extent to which isolation and negative test certification respectively reduce the risk of infection. To this end, a discrete-time SEIR model was used as the infectious disease model, and equations for calculating the conditional probability of non-infection status given negative test results on two different days were derived. Then the respective non-infection probabilities with two negative PCR test results, and with one negative PCR test result and one antigen test result, were quantified. By substituting initial parameters of the SEIR model into these probabilities, the present study revealed the following: (1) isolating close contact individuals can reduce by 80% the risk of infection during the first five days, but five more days are needed to reduce the risk 10% more, and seven more days to reduce the risk 20% more; and (2) if an individual with a negative PCR test result has a negative antigen test result the next day, then his or her infection probability is between 0.6% and 0.7%. Our results show that five-day isolation has a proportionally greater effect on risk reduction, compared to longer isolation; and thus, if an isolation period of longer than five days is contemplated, both the risk reduction and the negative effects from such increased isolation should be considered. Regarding negative test certification, our results provide those in managerial positions, who must decide whether to accept the risk and hold mass-gathering events, with quantitative information that may be useful in their decision-making.

## Introduction

Risk management of new coronavirus infections has become a significant public health issue. Isolation of symptomatic individuals was insufficient to control the spread of infection, because some infected people were asymptomatic, and the contagious period commenced roughly two days before symptoms appeared [8]. This latter may be somewhat shorter in the case of the omicron strain [10], but it still exists. Insufficient control when an infected individual appeared led to the global spread of the coronavirus disease of 2019 (COVID-19), in addition to the high reproduction number of COVID-19; for example, 2.5 [3] or 3.15 [7] for the wild-type strain, 7 for the delta strain [7], and greater still for the omicron strain [4].

As a countermeasure against infectious diseases, tests targeting individuals, such as polymerase chain reaction (PCR) and antigen tests, are widely used. Mina *et al*. [16] noted that a higher frequency of low-sensitivity antigen testing contributed more to infection control than a lower frequency of high-sensitivity PCR testing, and Du *et al*. [6] discussed strategies for effective testing systems, based on cost-effectiveness analysis of the tests and universal isolation. However, testing the entire public presents difficulties in terms of cost and test resources.

In contrast, it is a common strategy to test only those people who seem to be infected and may contribute to the infection spread, when an infected individual is confirmed. One strategy is to identify people retrospectively who have met infected people, and test and isolate them. In Japan, people who had highly infectious contact behavior with an individual infected with COVID-19, during the contagious period, are considered close contact people [17]. Here, infectious contact behavior means contact with an infected person, within 1 meter for more than 15 minutes, without wearing a mask. The contagious period is from two days before the onset of illness to when the infected person is isolated after diagnosis. Then, even if the contact person has negative test results, they are mandated to stay at home and be monitored for 14 days from the day after the last day of contact. It should be noted that, with the appearance of the omicron strain, the above periods have been revised from time to time; and in the latest situation, the monitoring period has been reduced from 14 to 7 days. While this strategy effectively prevents the spread of infection in advance, it can lead to a decline in socio-economic activity.

Another strategy is to proactively identify groups whose acts could contribute to the spread of infection and allow contact behavior for those with negative PCR- or antigen-test certification. Typical examples are easing restrictions for traveling and participating in mass-gathering events based on negative test certification. In the Japanese professional soccer league, for example, when an infected player or team staff member is confirmed, only those certified negative by an antigen test, which provides the result on the same day, are allowed to take part in the game [21, p12]. However, it is not fully clear how these negative test certifications contribute to infection reduction.

Vaccination is currently progressing worldwide, and strategies to allow socio-economic activity through vaccination or negative test certification are being discussed [20]. It is important, then, to evaluate how reasonable it is to shorten the isolation period through negative test certification, and how much risk reduction is associated with such certification.

The present study applied the SEIR (susceptible, exposed, infected, and recovered) compartmental model, a population dynamics model of infectious diseases, to address the issues above. There is a long history of research on mathematical models of infectious diseases. Kermack and McKendrick [13], the first study of this kind, proposed a SIR model, which incorporated the fact that epidemiological parameters change with the age of infection. Since then, theoretical studies involving infectious disease models have advanced considerably and the most fundamental concepts in infectious disease dynamics and evolutionary dynamics of infectious diseases, such as the definition of the primary reproduction number, have been formulated [5, 2]. These are the basis of current infectious disease research.

The present study, which is based on the SEIR model, simplifies the Kermack and McKendrick model and facilitates the interpretation of COVID-19 transmission. A number of studies have shown that this model can reproduce the characteristics of infectious disease dynamics [15, 6, 18]. However, it has been criticized for not accurately reproducing the dynamics of infectious diseases in large populations, such as entire countries; the main reason being that it assumes a homogeneous population, whereas real populations are composed of multiple heterogeneous subpopulations involving, for example, different social activities in different cases, or varying vaccination coverage in different regions. One of the study’s objectives, then, was to understand the general characteristics that apply to some extent in any population, and to provide information that will contribute to the management and control of infectious diseases in general, rather than simply replicating the dynamics of infectious diseases in one particular population. To this end, it was appropriate to utilize a simple model, and without restriction to the specificity of a given population. Although numerous studies have investigated specific populations, there have been very few attempts to provide such basic information.

The present study had two general aims: first, to investigate the reduction in infection risk with the use of negative test certification after an isolation period, and discuss possible reductions in the isolation period by combining the two most common tests; and second, to assess the extent to which negative antigen test results, without isolation, contribute to reducing the infection risk in a population that is likely to be infected. In addition, the present paper details the process of deriving the study’s results, and in concrete terms, so that readers can adapt the model to their specific cases.

## Methods

### Discrete SEIR Model

The present study applied a discrete-time Susceptible-Exposed-Infectious-Recovered (SEIR) model [9], in which individuals transition through five sequential statuses: susceptible (*S*), exposed (*E*), pre-symptomatic (*P*), infected (*I*), and recovered (*R*), with the infected status subdivided into symptomatic (*I*_*s*_) and asymptomatic (*I*_*a*_), in one of which the infected individual is registered.

The average duration of each state, in days, was adopted from He *et al*. [8], rounded to the nearest integer: after an individual is exposed to the coronavirus, they are in Status *E* for three days, Status *P* for two days, Status *I*_*s*_ or *I*_*a*_ for seven days, and then remain in Status *R* (Figure 1). Note that the first and second day of Status *P* are denoted by *P*_1_ and *P*_2_, respectively; and there is a possibility of being infected by COVID-19 two or more times, but this possibility is omitted in the present study.

**Figure 1:**
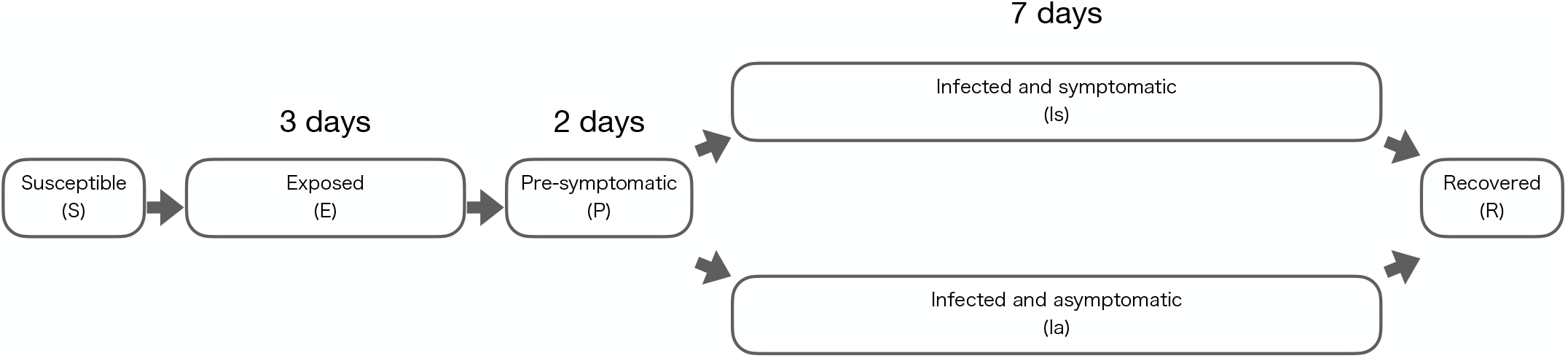
Discrete-time SEIR model used in the present study

Let *S*_0_, *E*_0_, *P*_1,0_, *P*_2,0_, and *I*_0_ be the initial populations of Statuses *S, E, P*_1_, *P*_2_, and *I*, respectively; and let day 0 be the initial day. Then the populations of day *k* (*k* = 1, 2, …, 14) can be easily calculated as in Table 1.

**Table 1:**
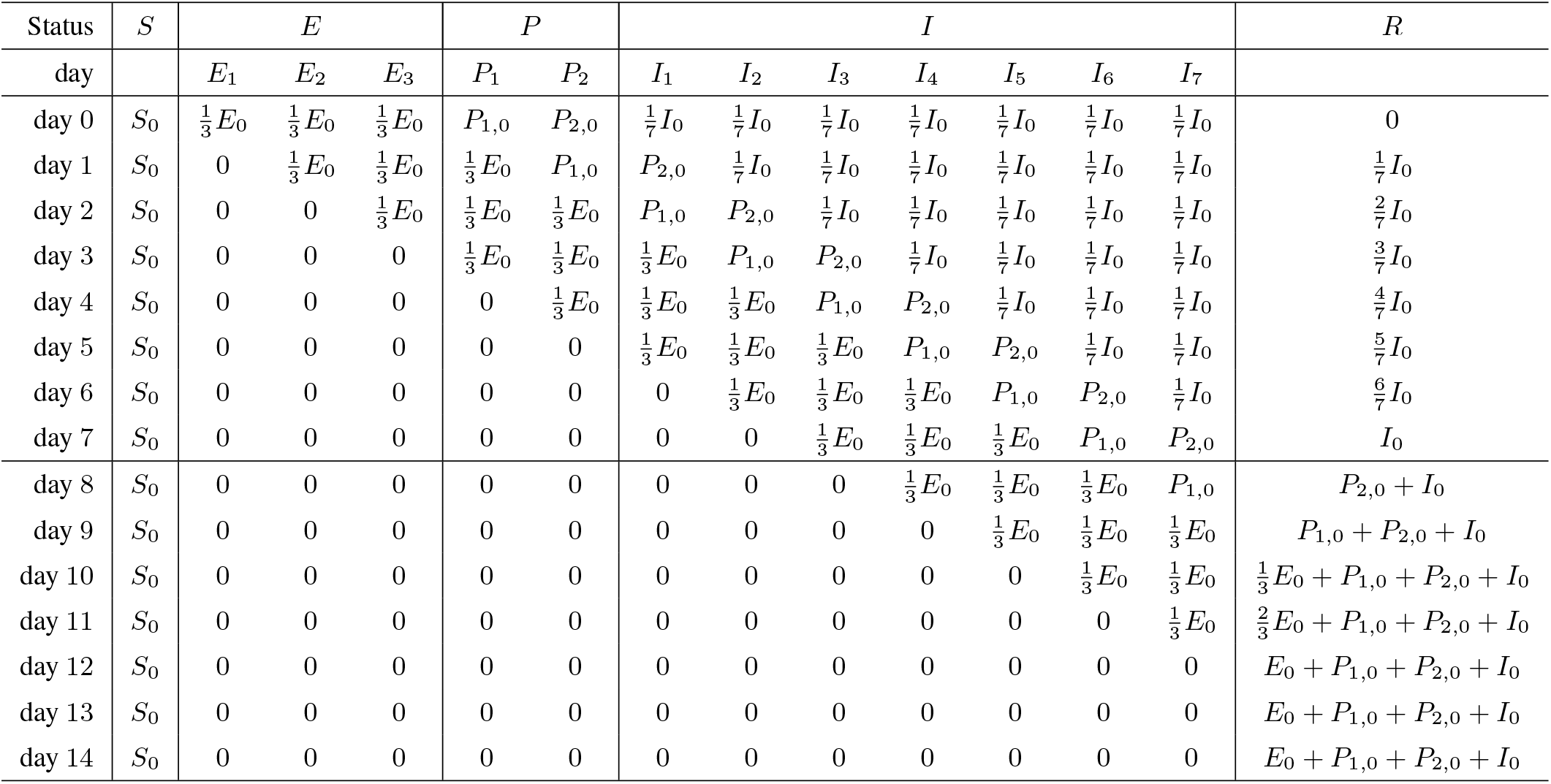
Time evolution of the distribution of people isolated on day 0, using the discrete SEIR model

### Two types of COVID19 tests

The present study deals with two types of tests (PCR and antigen tests) for determining whether an individual is infected with COVID-19. Kucirka *et al*. [14] and Brümmer *et al*. [**?**] reported that the sensitivity depended on an individual’s status. Let *a*_1_, *a*_2_, and *a*_*I*_, then, be sensitivities of COVID-19 PCR tests for individual status *P*_1_, *P*_2_, and *I*, respectively. Then *a*_1_ = 0.33, *a*_2_ = 0.67, and *a*_*I*_ = 0.80. Further, let *b* be the specificity of a COVID-19 PCR test for individual Statuses *S* and *E*, then *b* = 0.999. Table 2 summarizes the sensitivity and specificity of COVID-19 PCR tests.

**Table 2:**
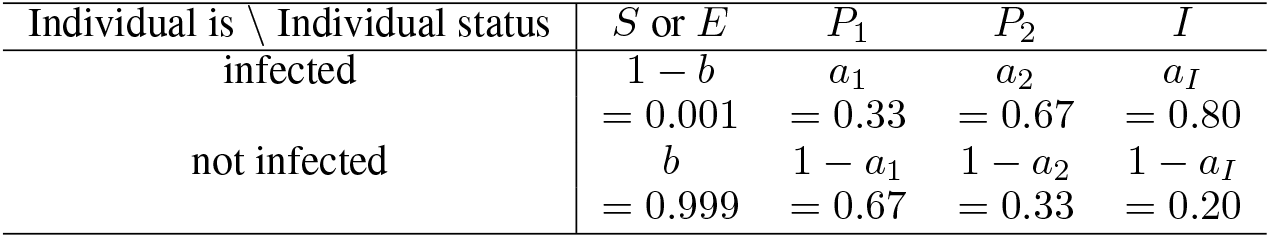
Accuracy of COVID-19 PCR tests

COVID-19 antigen tests have 0.7-times the sensitivity and the same specificity as PCR tests [**?**]. In other words, the sensitivities of antigen tests for individual Statuses *P*_1_, *P*_2_, and *I* are 0.7*a*_1_ = 0.23, 0.7*a*_2_ = 0.47, and 0.7*a*_*I*_ = 0.56, respectively; while the specificity for individual Statuses *S* and *E* is *b* = 0.999. Table 3 summarizes the sensitivity and specificity of COVID-19 antigen tests.

**Table 3:** Accuracy of COVID-19 antigen tests

| Individual is \ Individual status | $S$ or $E$ | $P_1$ | $P_2$ | $I$ |
| --- | --- | --- | --- | --- |
| infected | $1 - b$<br>= 0.001 | $0.7a_1$<br>= 0.23 | $0.7a_2$<br>= 0.47 | $0.7a_I$<br>= 0.56 |
| not infected | $b$<br>= 0.999 | $1 - 0.7a_1$<br>= 0.77 | $1 - 0.7a_2$<br>= 0.53 | $1 - 0.7a_I$<br>= 0.44 |

It should be remarked that the sensitivity of antigen tests varies with the type of test, and can be less than 0.7 [19]. Further, in the case of the omicron strain, one study has reported that the sensitivity of nasal antigen tests is low within three days from onset [1].

### Conditional probability of non-infection status given negative test results

Hereafter, let ⊕_*k*_ and ⊖_*k*_ be situations in which an individual has a positive and a negative PCR test result, respectively, on day *k* (*i* = 0, …, 14); and *P*(*X*|*Y*) represents the probability that the individual is in Status *X* in Situation *Y*, where *X* is one of *S, E, P*_1_, *P*_2_, *I, R*, or their union, and *Y* is one of ⊕_*k*_, ⊖_*k*_, or their intersection.

### Conditional probability of non-infection status given one negative PCR test result

This clause derives the non-infected probability when an individual has a negative test result on day 0. In other words, *P*(*S*_0_ ∪ *R*_0_|⊖_0_) is derived. Note that cursive script *P* stands for the probability, while capital letter *P* stands for an individual’s status. Let 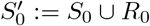; that is, 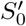 means a non-infection status on day 0. Since the overall status on day 0, say Ω_0_, equals *S*_0_ ∪ *E*_0_ ∪ *P*_1,0_ ∪ *P*_2,0_ ∪ *I*_0_ ∪ *R*_0_, the following holds from Bayes’ theorem.

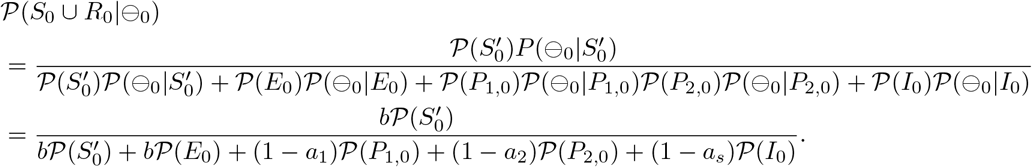

### Conditional probability of non-infected status given two negative PCR test results

This clause derives the non-infection probability when an individual has a negative PCR test result on both day 0 and day *k k* (1 ≤ *k* ≤ 14). In other words, *P*(*S*_*k*_ ∪ *R*_*k*_| ⊖_0_ ∩⊖_*k*_) is derived. Let 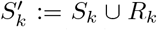; that is, 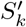 indicates non-infection status at day *k* (0 ≤ *k* ≤ 14). First, from Bayes’ theorem, the following holds.

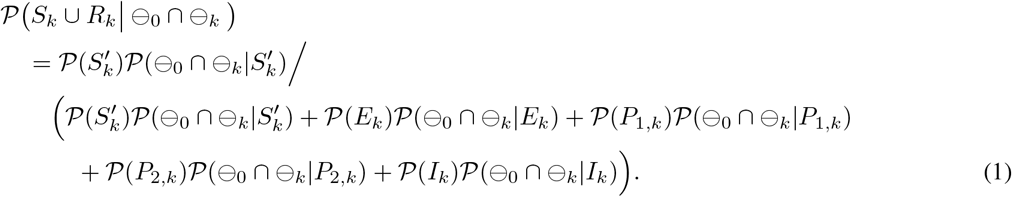

Next, in order to calculate (1), 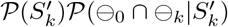 is derived. By the definitions of the conditional probability and Ω_0_,

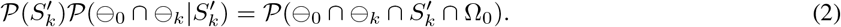

Then, from De Morgan’s laws, the following holds.

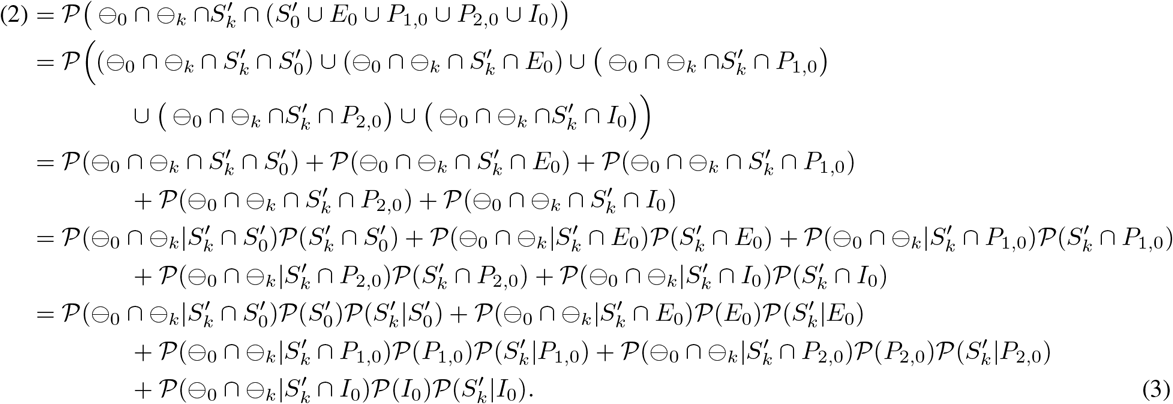

In the same way, the following hold.

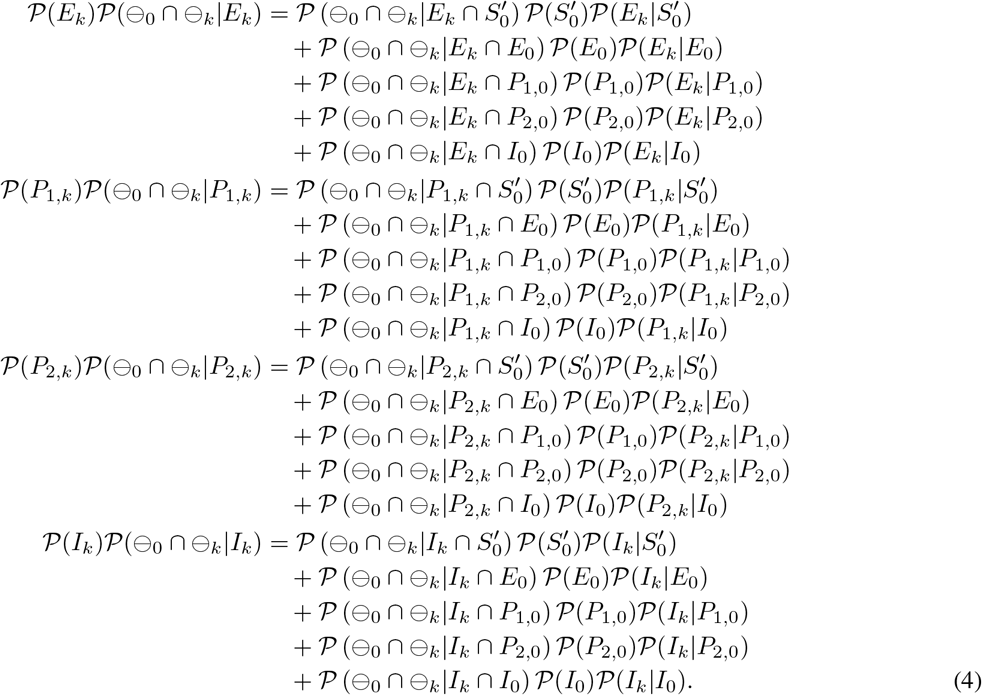

Finally, by substituting (3)–(4) into (1), the desired probability can be derived.

## Results

### Non-infection probability under a negative PCR test result on both day 0 and day *k*

#### The case where *k* = 1

From Table 1, the following relationships between the day 0 and day 1 statuses hold.

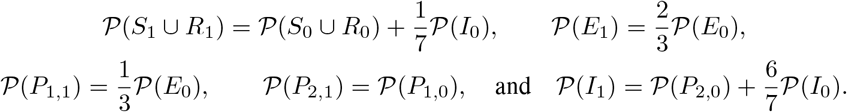

Therefore, the members on the right-hand side of (1) can be calculated as follows.

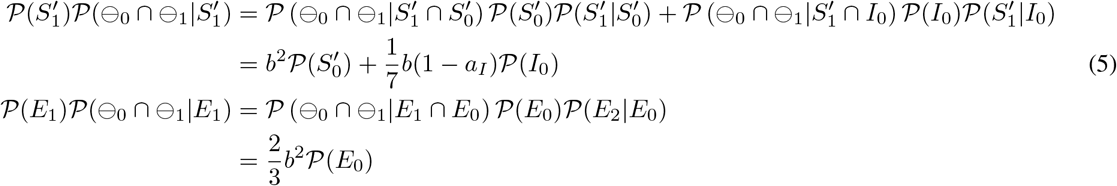

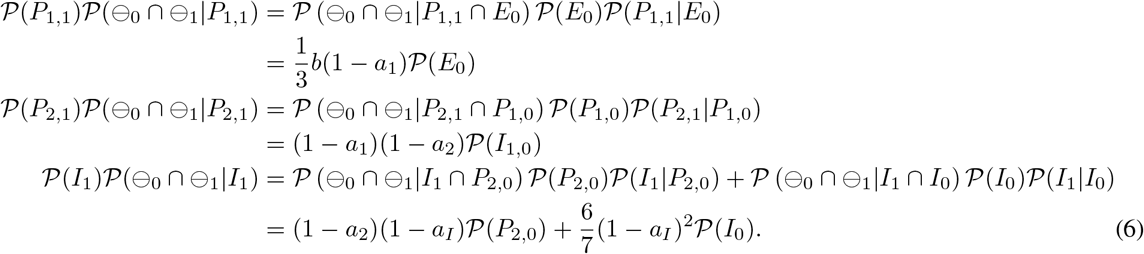

By substituting (5)–(6) and *k* = 1 into (1), the desired probability can be derived as follows.

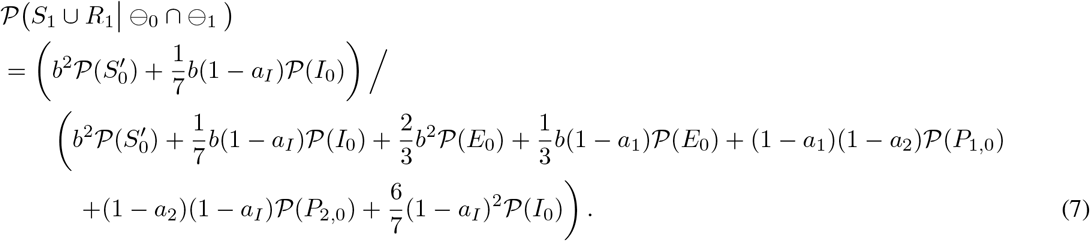

In the same way, the non-infection probability for an individual with a negative PCR test result on both day 0 and day *k* for any *k*(= 2, …, 14) can be derived. Only the results are shown in the following clauses.

#### The case where *k* = 2

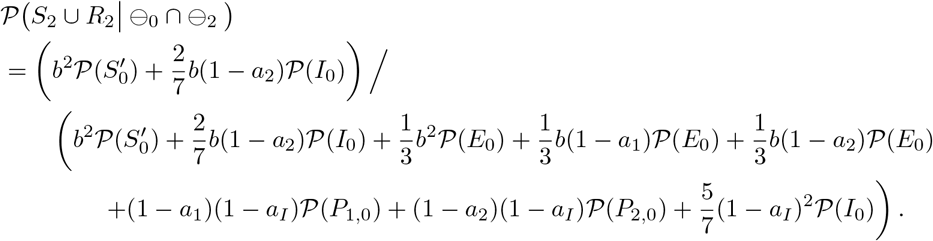

#### The case where *k* = 3

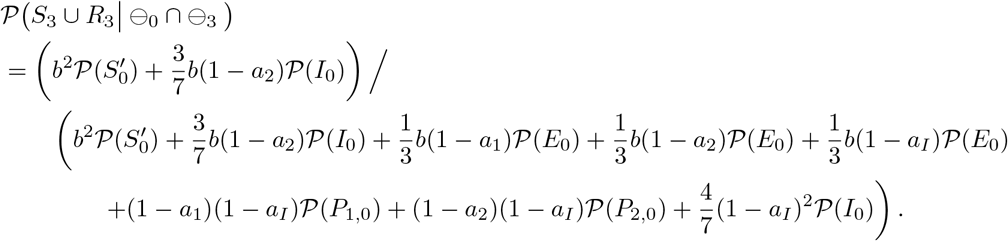

#### The case where *k* = 4

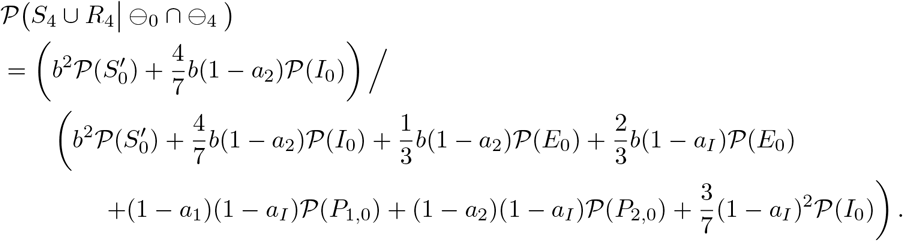

#### The case where *k* = 5

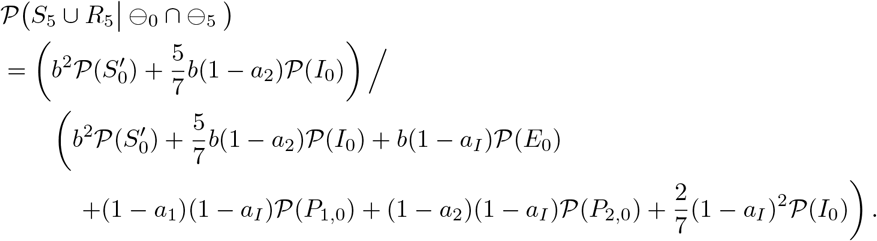

#### The case where *k* = 6

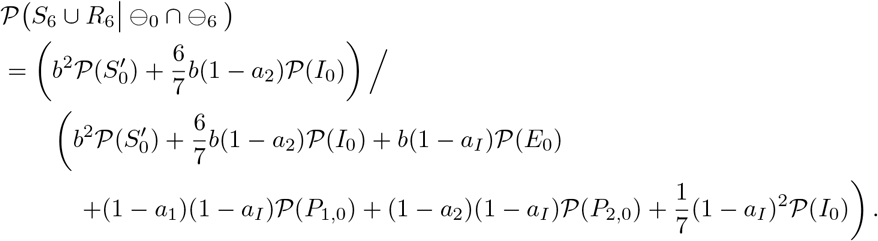

#### The case where *k* = 7

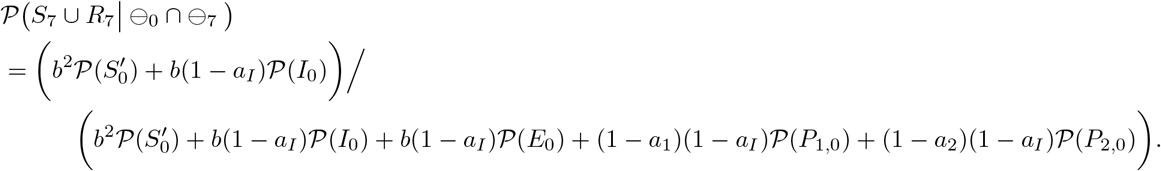

#### The case where *k* = 8

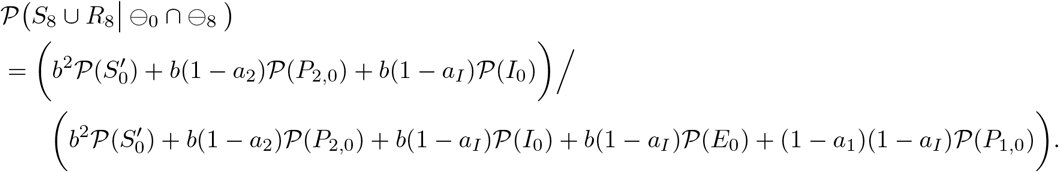

#### The case where *k* = 9

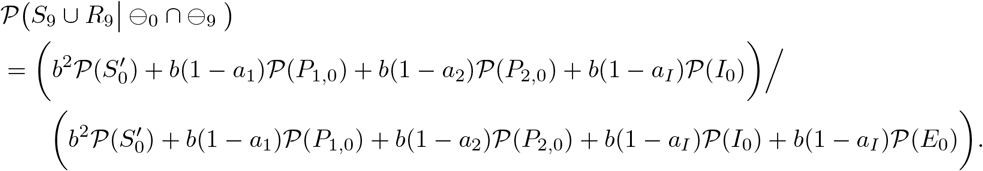

#### The case where *k* = 10

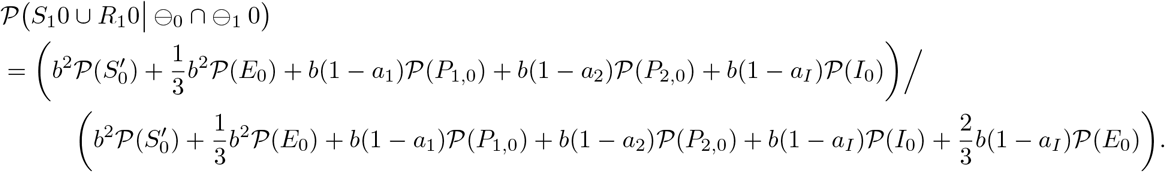

#### The case where *k* = 11

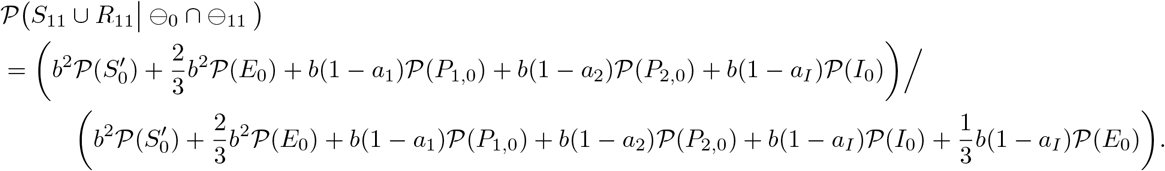

#### The case where *k* = 12, 13, and 14

Since every individual is in Status *R* after day 12, *P*(*S*_12_ ∪ *R*_12_| ⊖_0_ ∩ ⊖_12_), *P*(*S*_13_ ∪ *R*_13_| ⊖_0_ ∩ ⊖_13_), and *P*(*S*_14_ ∪ *R*_14_ | ⊖_0_ ∩ ⊖_14_) are identical.

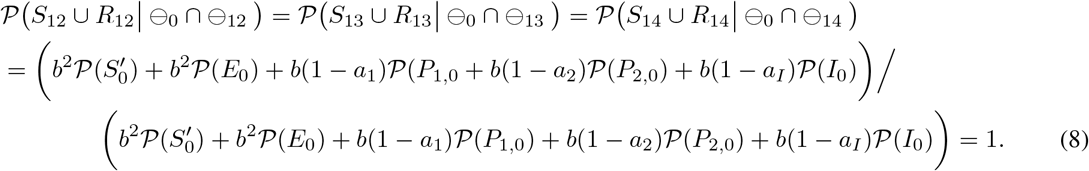

### Non-infection probability with one negative PCR test result and one negative antigen test result

Since the sensitivity of COVID-19 antigen tests are 0.7 times that of COVID-19 PCR tests, we can obtain the probability 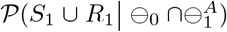 in a manner similar to the derivation of (7), but with 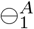 indicating that an individual has a negative antigen test result on day 1.

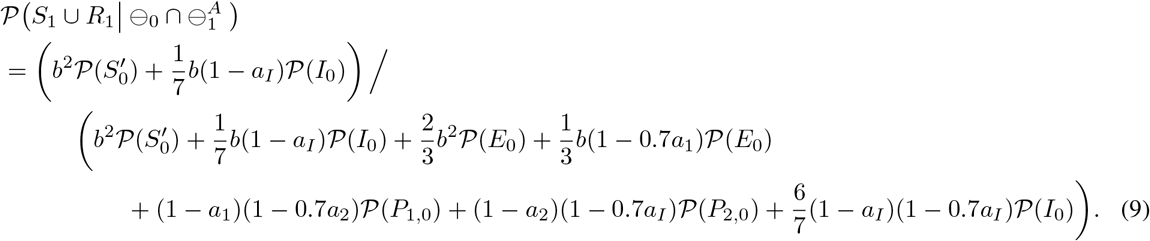

### Infection probability for isolated people

This subsection demonstrates the respective infection probabilities for the following two-types of close contact people after a certain isolation period:

i. isolated when an individual with a positive PCR test result appears in the community;
ii. isolated when a symptomatic individual appears.

In order to demonstrate numerical results, the present study applied the initial-population parameters in Kamo *et al*. [12]. For case (i),

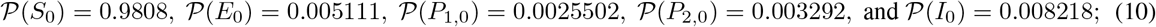

while, for case (ii),

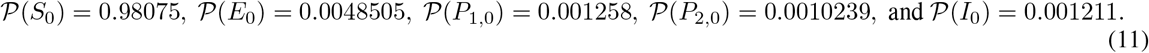

By substituting *a*_1_, *a*_2_, *a*_*I*_, and (10) or (11) into (7)–(8), and subtracting them from 1, we obtain estimates of the infection probabilities on day *k* for an individual with a negative PCR test result on both day 0 and day *k*. Table 4 shows these probabilities, and Figure 2 presents the same information using a scatter plot. In the graph, for *x* ≠ 0, the *x*-axis represents the day when an individual takes the second PCR test, and the *y*-axis the infection probability for an individual with a negative PCR test result on both day 0 and day *x*. For *x* = 0, the corresponding *y* value represents the infection probability for an individual with a negative PCR test result on day 0. The dots show the infection probabilities for Case (i), and the squares for Case (ii). The solid and dashed lines connect the dots and squares, respectively.

**Table 4:** List of infection probabilities on day *k* for an individual with a negative PCR test result on both day 0 and day *k*. Note that the results at *k* = 0 indicate the probabilities on day 0 for an individual with a negative PCR test result on day 0.

| $k$ | Case(i) | Case(ii) |
| --- | --- | --- |
| 0 | 0.009811 | 0.008602 |
| 1 | 0.005810 | 0.005205 |
| 2 | 0.004386 | 0.003968 |
| 3 | 0.002959 | 0.002588 |
| 4 | 0.002098 | 0.001746 |
| 5 | 0.001739 | 0.001379 |
| 6 | 0.001690 | 0.001308 |
| 7 | 0.001642 | 0.001237 |
| 8 | 0.001386 | 0.001158 |
| 9 | 0.001037 | 0.0009855 |
| 10 | 0.0006905 | 0.0006561 |
| 11 | 0.0003448 | 0.0003276 |
| 12 | 0 | 0 |
| 13 | 0 | 0 |
| 14 | 0 | 0 |

**Figure 2:**
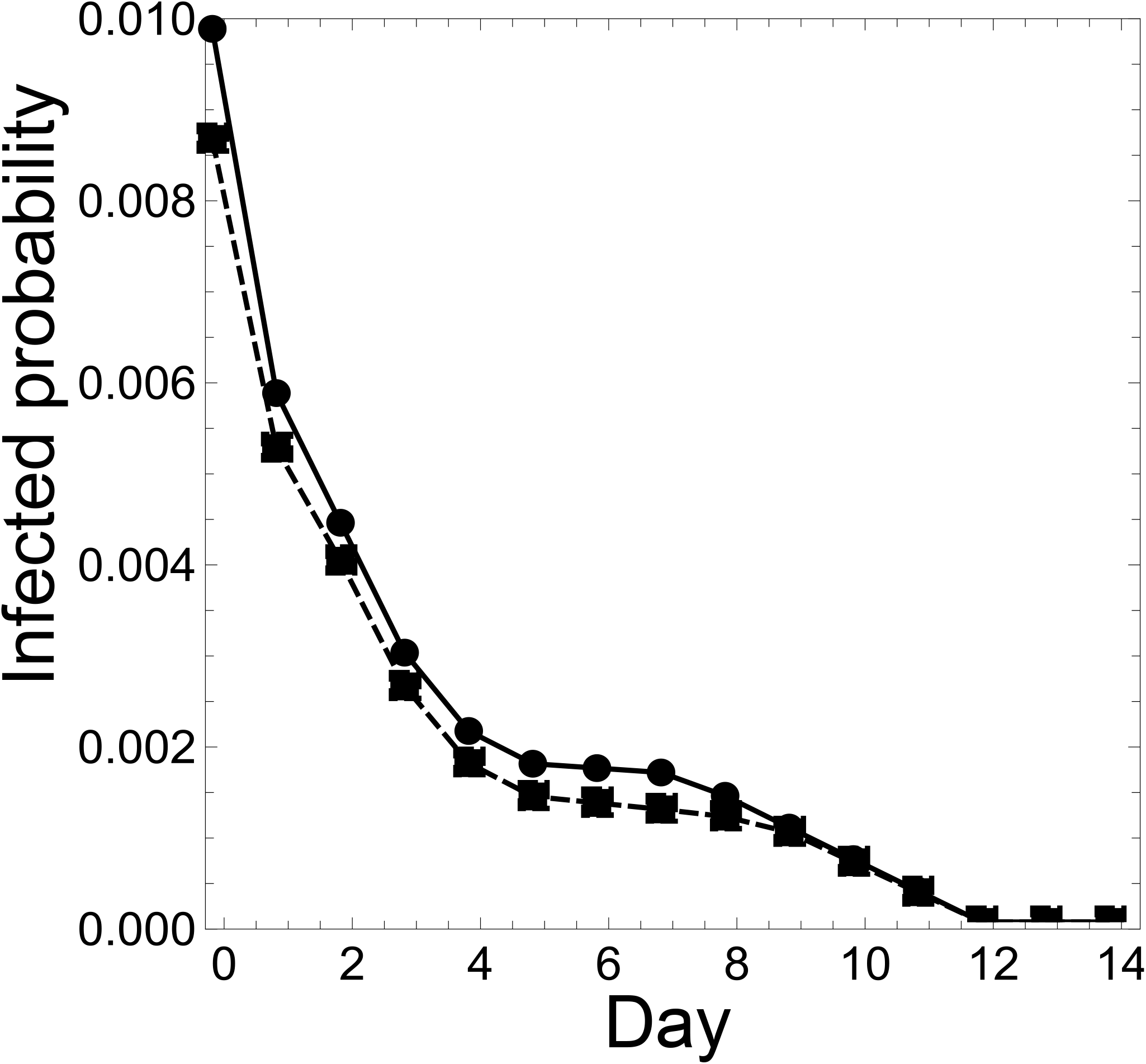
For *x* ≠ 0, the *x*-axis represents the day when an individual takes the second PCR test, and the *y*-axis the infection probability for an individual with a negative PCR test result on both day 0 and day *x*. For *x* = 0, the corresponding *y* value represents the infection probability for an individual with a negative PCR test result on day 0. The dots show the infection probabilities for Case (i), and the squares for Case (ii). The solid and dashed lines connect the dots and squares, respectively.

### Infection probability for close contact people

#### Infection probability for close contact people with two negative PCR tests

This clause assumes that 0.25 of the total population are close contact people, and *p* of the total number of infected people are close contact people. Then the initial-population parameters are, for Case (i),

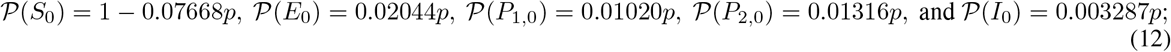

while, for Case (ii),

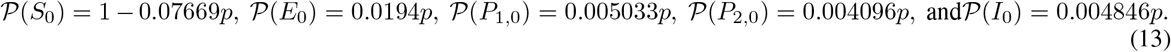

When the proportion *p* is varied, Table 5 shows the initial populations of close contact people who are isolated at the appearance of an individual with a positive PCR test result (Case (i-Cls)) or who is symptomatic (Case (ii-Cls)).

**Table 5:** Initial populations of close contact people who are isolated at the appearance of an individual with a positive PCR test result (Case (i-Cls)) or who is symptomatic (Case (ii-Cls), with various percentages of infected people in the close contact group (*p*).

| $p$ | Case | $P(S_0)$ | $P(E_0)$ | $P(Ia_{1,0})$ | $P(Ia_{2,0})$ | $P(I_0)$ |
| --- | --- | --- | --- | --- | --- | --- |
| $p = 0.25$ | Case(i-ClS) | 0.9808 | 0.005111 | 0.002550 | 0.003291 | 0.008218 |
|  | Case(ii-ClS) | 0.9808 | 0.004851 | 0.001258 | 0.001024 | 0.01211 |
| $p = 0.30$ | Case(i-ClS) | 0.9770 | 0.006133 | 0.00306 | 0.003949 | 0.009862 |
|  | Case(ii-ClS) | 0.9769 | 0.005821 | 0.001510 | 0.001229 | 0.01454 |
| $p = 0.50$ | Case(i-ClS) | 0.9617 | 0.01022 | 0.005100 | 0.006582 | 0.01644 |
|  | Case(ii-ClS) | 0.9615 | 0.009701 | 0.002517 | 0.002048 | 0.02423 |
| $p = 0.60$ | Case(i-ClS) | 0.9540 | 0.01227 | 0.00612 | 0.007899 | 0.01972 |
|  | Case(ii-ClS) | 0.9538 | 0.01164 | 0.003020 | 0.002457 | 0.02907 |
| $p = 0.90$ | Case(i-ClS) | 0.9310 | 0.01840 | 0.009181 | 0.01185 | 0.02959 |
|  | Case(ii-ClS) | 0.9307 | 0.01746 | 0.004530 | 0.003686 | 0.04361 |

By substituting *a*_1_, *a*_2_, *a*_*I*_, and (12) or (13) into (7)–(8), and subtracting them from 1, we obtain estimates of the infection probabilities on day *k* for an isolated individual with a negative PCR test result on both day 0 and day *k*, for the cases where *p* = 0.25, 0.30, 0.50, 0.60, and 0.90. Tables 6 and 7 show these infection probabilities for Case (i-Cls) and Case (ii-Cls), respectively; and Figures 3 and 4 present the same information using scatter plots. In these graphs, the *x*-axes represent the day where an individual takes the second PCR test (except *x* = 0), while the *y*-axes represent the infection probability (in the case of *x* = 0, the infection probability for an individual with a negative PCR test result on day 0).

**Table 6:** Infection probability for an individual in Case (i-Cls) with a negative PCR test result on both day 0 and day *k* (*k* ≥ 1), under the assumption that the percentage of infected people in the close contact people is *p*. When *k* = 0, the table shows the infection probability for an individual in Case (i-Cls) with a negative PCR test result on day 0.

| $k \setminus p$ | Case(i-ClS) | | | | |
| --- | --- | --- | --- | --- | --- |
| | $p = 0.25$ | $p = 0.30$ | $p = 0.50$ | $p = 0.60$ | $p = 0.90$ |
| 0 | 0.009811 | 0.01180 | 0.01981 | 0.02387 | 0.03622 |
| 1 | 0.005810 | 0.006991 | 0.01178 | 0.01421 | 0.02167 |
| 2 | 0.004386 | 0.005278 | 0.008901 | 0.01074 | 0.01641 |
| 3 | 0.002959 | 0.003562 | 0.006013 | 0.007262 | 0.01111 |
| 4 | 0.002098 | 0.002526 | 0.004267 | 0.005154 | 0.007891 |
| 5 | 0.001739 | 0.002093 | 0.003536 | 0.004272 | 0.006541 |
| 6 | 0.001690 | 0.002035 | 0.003437 | 0.004152 | 0.006356 |
| 7 | 0.001642 | 0.001977 | 0.003339 | 0.004033 | 0.006172 |
| 8 | 0.001386 | 0.001669 | 0.002815 | 0.003399 | 0.005195 |
| 9 | 0.001037 | 0.001248 | 0.002103 | 0.002538 | 0.003872 |
| 10 | 0.0006905 | 0.0008307 | 0.001398 | 0.001686 | 0.002568 |
| 11 | 0.0003448 | 0.0004147 | 0.0006972 | 0.0008404 | 0.001278 |
| 12 | 0 | 0 | 0 | 0 | 0 |
| 13 | 0 | 0 | 0 | 0 | 0 |
| 14 | 0 | 0 | 0 | 0 | 0 |

**Table 7:** Infection probability for an individual in Case (ii-Cls) with a negative PCR test result on both day 0 and day *k* (*k* ≥ 1), under the assumption that the percentage of infected people in the close contact people is *p*. When *k* = 0, the table shows the infection probability for an individual in Case (ii-Cls) with a negative PCR test result on day 0.

| $k \setminus p$ | Case(ii-CIs) | | | | |
| --- | --- | --- | --- | --- | --- |
| | $p = 0.25$ | $p = 0.30$ | $p = 0.50$ | $p = 0.60$ | $p = 0.90$ |
| 0 | 0.008602 | 0.01034 | 0.01739 | 0.02096 | 0.03187 |
| 1 | 0.005205 | 0.006263 | 0.01056 | 0.01274 | 0.01944 |
| 2 | 0.003968 | 0.004776 | 0.008056 | 0.009726 | 0.01486 |
| 3 | 0.002588 | 0.003116 | 0.005260 | 0.006353 | 0.009719 |
| 4 | 0.001746 | 0.002102 | 0.003549 | 0.004288 | 0.006564 |
| 5 | 0.001379 | 0.001660 | 0.002804 | 0.003388 | 0.005186 |
| 6 | 0.001308 | 0.001575 | 0.002659 | 0.003212 | 0.004916 |
| 7 | 0.001237 | 0.001490 | 0.002514 | 0.003037 | 0.004646 |
| 8 | 0.001158 | 0.001394 | 0.002352 | 0.002840 | 0.004343 |
| 9 | 0.0009855 | 0.001186 | 0.002001 | 0.002415 | 0.003690 |
| 10 | 0.0006561 | 0.0007895 | 0.001330 | 0.001605 | 0.002448 |
| 11 | 0.0003276 | 0.0003941 | 0.0006633 | 0.0008000 | 0.001218 |
| 12 | 0 | 0 | 0 | 0 | 0 |
| 13 | 0 | 0 | 0 | 0 | 0 |
| 14 | 0 | 0 | 0 | 0 | 0 |

**Figure 3:**
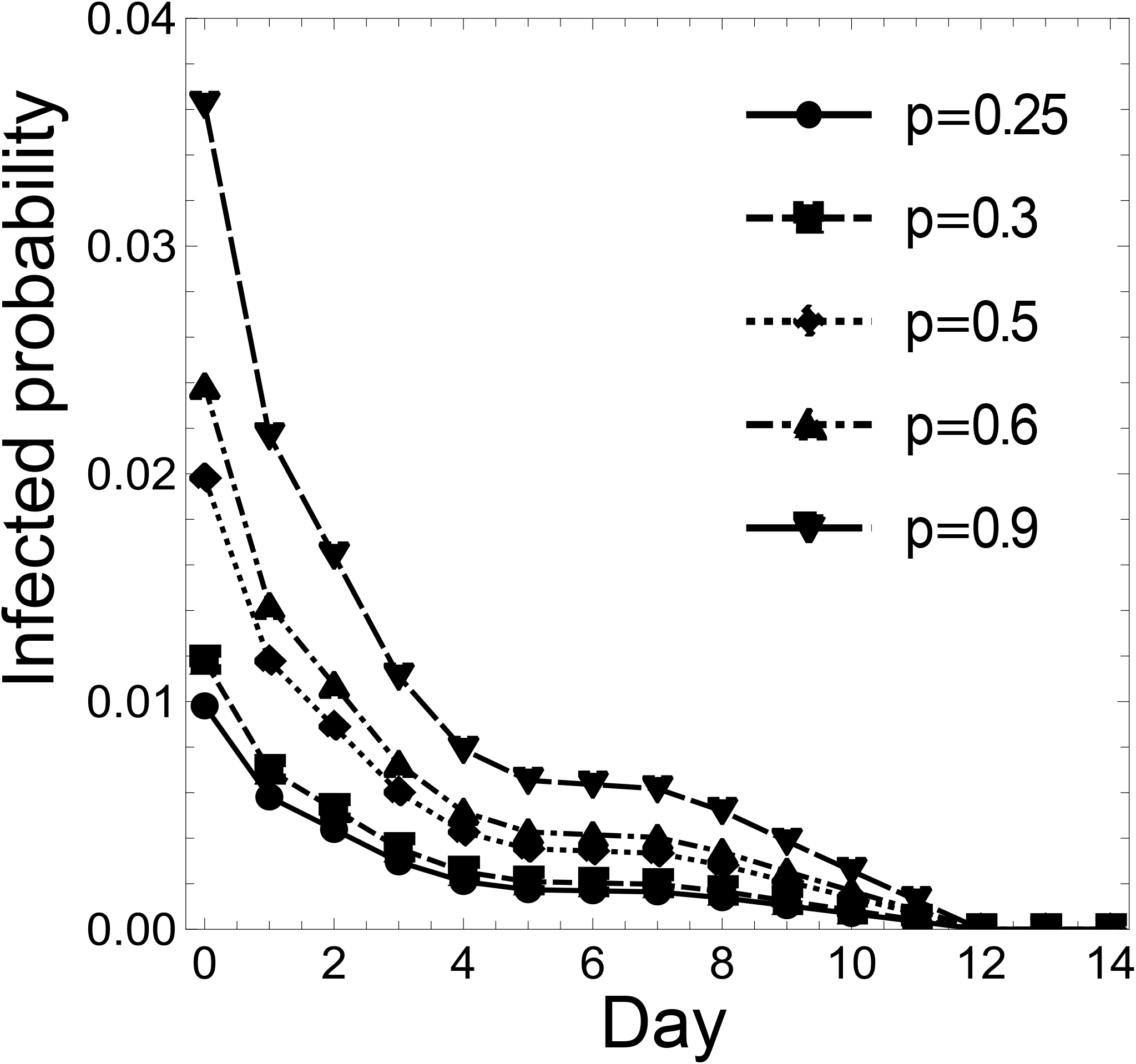
For *x* ≠ 0, the *x*-axis represents the day when an individual takes the second PCR test, and the *y*-axis the infection probability for an individual with a negative PCR test result on both day 0 and day *x*, for Case (i-Cls). For *x* = 0, the corresponding y value represents the infection probability for an individual with a negative PCR test result on day 0, for Case (i-Cls). The dots, squares, diamonds, upward triangles, and downward triangles show the infection probabilities for *p* = 0.25, 0.30, 0.50, 0.60, 0.90, respectively, for Case (i-Cls); and the solid, short-dashed, dotted, dash-dotted, and long-dashed lines connect the dots, squares, diamonds, upward triangles, and downward triangles, respectively.

**Figure 4:**
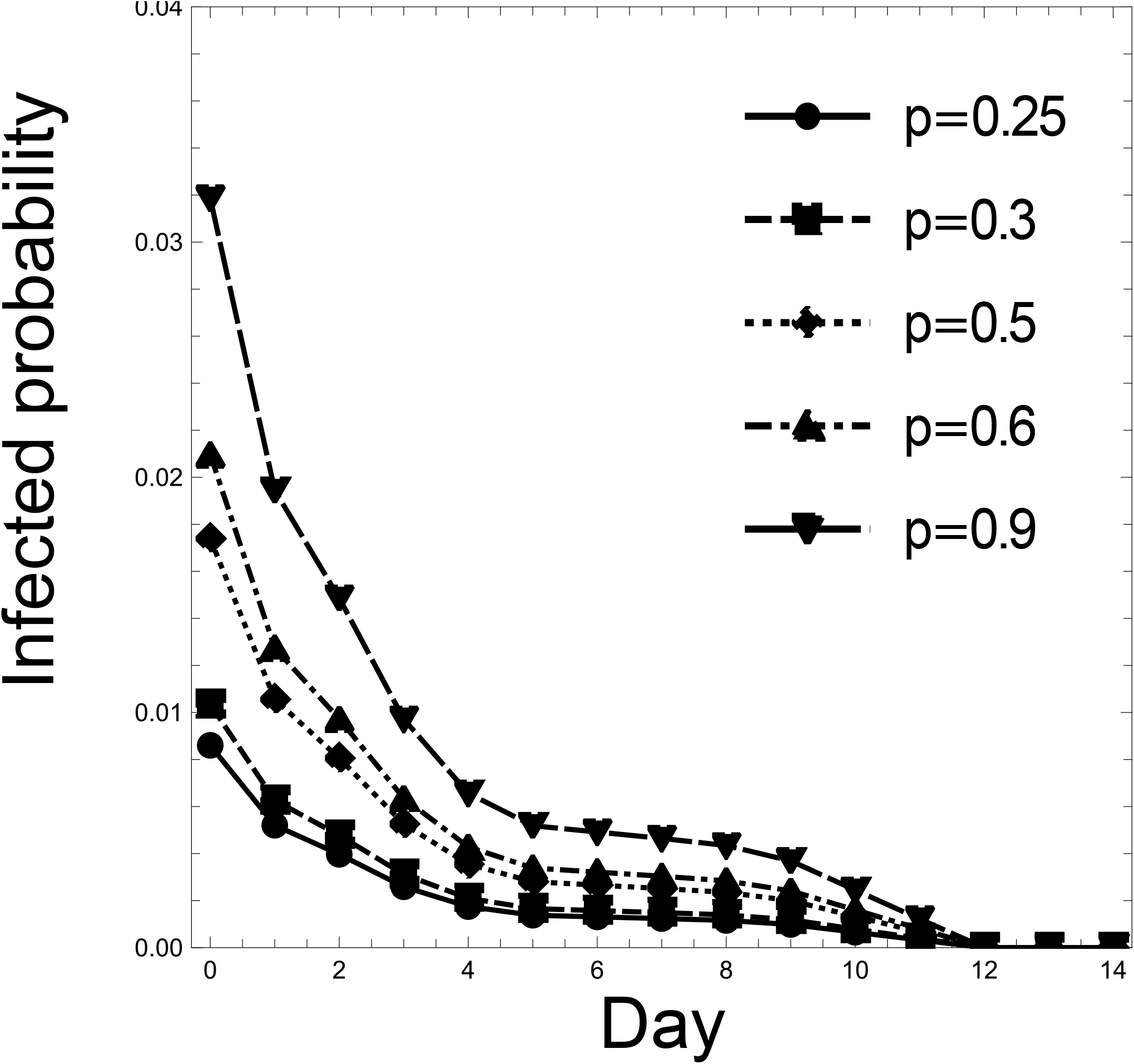
For *x* ≠ 0, the *x*-axis represents the day when an individual takes the second PCR test, and the *y*-axis the infection probability for an individual with a negative PCR test result on both day 0 and day *x*, for Case (ii-Cls). For *x* = 0, the corresponding y value represents the infection probability for an individual with a negative PCR test result on day 0, for Case (ii-Cls). The dots, squares, diamonds, upward triangles, and downward triangles show the infection probabilities for *p* = 0.25, 0.30, 0.50, 0.60, 0.90, respectively, for Case (ii-Cls); and the solid, short-dashed, dotted, dash-dotted, and long-dashed lines connect the dots, squares, diamonds, upward triangles, and downward triangles, respectively.

### Infection probability for close contact people with one negative PCR test and one negative antigen test

Using the same conditions as in the previous clause, this clause deals with the situation where an individual takes a COVID-19 antigen test on day 1. Also, for each *p*, the infection probabilities for people with a negative PCR test result on day 1 and a negative antigen test result on day 0 are compared to those for people with negative PCR test results on both day 0 and day 1.

By substituting *a*_1_, *a*_2_, *a*_*I*_, and (10) or (11) into (9), and subtracting it from 1, we obtain estimates of the infection probabilities on day 1 for an individual with a negative PCR test result on day 0 and a negative antigen test result on day 1. Table 8 shows these infection probabilities and the comparison with the case where an individual has negative PCR test results on both day 0 and day 1. In the table, Δ stands for the increment in the infection probability when the test type on day 1 is changed from a PCR to an antigen test.

**Table 8:** Infection probability for an individual with a negative PCR test result on day 0 and a negative PCR or antigen test result on day 1. The symbol Δ represents the difference in probability.

|  | Case(i) | Case(ii) |
| --- | --- | --- |
| PCR test at day 1 | 0.005810 | 0.005205 |
| Antigen test at day 1 | 0.006944 | 0.006122 |
| $\Delta$ | +0.001134 | +0.0009170 |

By substituting *a*_1_, *a*_2_, *a*_*I*_, and (12) or (13) into (9), and subtracting it from 1, we obtain estimates of the infection probabilities on day 1 for an isolated individual with a negative PCR test result on day 0 and a negative antigen test result on day 1, for the cases where *p* = 0.25, 0.30, 0.50, 0.60, and 0.90. Table 9 shows these infection probabilities and the comparison with the case where an individual has a negative PCR test result on both day 0 and day 1, for both Case (i-Cls) and Case (ii-Cls). In the table, Δ stands for a similar increment as in Table 8.

**Table 9:** Infection probability for an individual with a negative PCR test result on day 0 and a negative PCR or antigen test result on day 1. The symbol Δ represents the difference in probability, under the assumption the percentage of infected people in the close contact people is *p*.

| $p$ | Test type at day 1 | Case(i-CIs) | Case(ii-CIs) |
| --- | --- | --- | --- |
| $p = 0.25$ | PCR | 0.005810 | 0.005205 |
|  | Antigen test | 0.006944 | 0.006122 |
| | $\Delta$ | +0.001134 | +0.0009170 |
| $p = 0.30$ | PCR | 0.0069961 | 0.006263 |
|  | Antigen | 0.08353 | 0.007366 |
| | $\Delta$ | +0.001136 | +0.001103 |
| $p = 0.50$ | PCR test | 0.01178 | 0.01056 |
|  | Antigen test | 0.01406 | 0.01241 |
| | $\Delta$ | +0.00228 | +0.00185 |
| $p = 0.60$ | PCR test | 0.01421 | 0.01274 |
|  | Antigen test | 0.01696 | 0.01497 |
| | $\Delta$ | +0.00275 | +0.00223 |
| $p = 0.90$ | PCR test | 0.02167 | 0.01944 |
|  | Antigen test | 0.02582 | 0.02281 |
| | $\Delta$ | +0.00415 | +0.00337 |

## Discussion

### Reduction of the isolation period

As noted in the Introduction, in Japan, close contact people previously had to be isolated for 14 days from the last day of contact, but this period has been reduced from 14 to 7 days with the appearance of the omicron strain.

Figure 2 shows that the infection probability decreases sharply from day 0 to day 5, remains roughly the same from day 6 to day 8, and then decreases from day 9 to day 12, for both Case (i) and Case (ii). This tendency implies that a five-day isolation period could be chosen to reduce the isolation period with less increased risk. Reducing the isolation period from 14 to 5 days corresponds to allowing for the risk of a 0.17% and a 0.14% increase in the infection probability for Case (i) and Case (ii), respectively. In other words, the expected number of infected people in a group of 1, 000 people would be less than two for both cases. Reducing the period from 7 to 5 days corresponds to the risk of a roughly 0.01% increase in the infection probability for both cases, meaning that the expected number of infected people in a group of 10, 000 people would be roughly one. These results suggest that if we consider a small group, the increased risk would be negligible in these cases.

Next, when close contact people constitute 25% of a given group, Figures 3 and 4 show the same pattern as Figure 2. Therefore, here too, 5 days are a candidate for a reduced isolation period. From Table 6, the reduction in Case (i-Cls) corresponds to allowing for the risk of a 0.17%, 0.21%, 0.35%, 0.43%, and 0.65% increase in the infection probability for *p* = 0.25, 0.30, 0.50, 0.60, and 0.90, respectively. From Table 7, the reduction in Case (ii-Cls) corresponds to allowing for the risk of a 0.14%, 0.17%, 0.28%, 0.34%, and 0.52% increase in the infection probability for *p* = 0.25, 0.30, 0.50, 0.60, and 0.90, respectively. As the infection probability on day 5 is less than 20% of that on day 0, the five-day isolation period and double test would result in an 80% risk reduction. In sum, the maximum expected number of infected people in a group of 1, 000 people is less than 7 in every case. If one wants to achieve 90% and 100% risk reduction, the isolation period should be 10 and 12 days, respectively. This implies that we can eliminate 80% of the risk during the first 5 days but need 5 more days to eliminate 10% more and 7 more days to eliminate 20% more.

In summary, a five-day isolation period has a significantly greater per diem effect on risk reduction than longer isolation. Thus, if an isolation period of longer than five days is contemplated, both the risk reduction and the negative effects of such prolonged isolation should be considered; and in the case of the omicron strain, an even shorter period may be sufficient to reduce the risk, because the incubation period is shorter.

### Effect of negative test certification

Consider a team, such as a Japanese professional soccer or baseball team, and the situation where

- all the members of the team take PCR tests on the day before a game;
- an individual with a positive test result appears, but the other individuals have negative test results; and
- the other individuals take antigen tests on game day, and individuals with negative antigen test results are allowed to participate in the game.

By applying the study’s results for day 1, we can discuss the effect of negative test certification in the above situation. Table 8 shows that the infection probability for an individual is between 0.6% and 0.7%; that is, the infection probability for an individual with negative antigen test certification is very low. For example, when 30 people with negative antigen test certification take part in a game, the probability that at least one of them is infected is about 20% because 1 − (1 − 0.006)^30^ = 0.17 and 1 − (1 − 0.007)^30^ = 0.19. It is up to the manager to decide whether to accept this risk and continue with the game, but our results will help such officials, by providing them with such quantitative information, which may be useful for their decision-making. If PCR tests are used instead of antigen tests, the expected infection probability decreases by only about 0.1%; that is, there is little difference between the two tests in this situation.

It should be noted that our model assumes that the people in a given group are isolated until day 1, but they are not isolated strictly from day 0 to day 1 in the above scenario. However, people’s behavior can typically be well managed in the case of small groups, which means that the probability of a new infection appearing in that one day is almost negligible. Our results show that administering an antigen test on game day can be useful for determining whether a game should be held, and which team members should be allowed to participate in the game, when an individual with a positive test result appears before the game. Further, this serves as an example of how inexpensive and rapid antigen tests can be utilized effectively to enable commercial activities and avoid economic shutdowns.

### Applicability and limitations of the present study

We conclude the section by discussing the applicability and limitations of the study. First, it should be noted that the study results were generated by a simple discrete-time SEIR model with averaged parameters based on the coronavirus information as of 2020; thus, the estimated infection probabilities and risk magnitudes are also averages as of 2020. If one wished to be especially careful, focusing on the 95th percentile or a sensitive population for example, the parameters should be reconsidered. Further, since viruses typically mutate, the parameters must be adjusted according to the latest situation. In particular, in the case of the omicron strain, it is important to utilize the latest data, as new information of the sort required to perform the same analysis as in the present study is being reported daily. Equations (5)–(9) can be used for such updates.

Second, the discussion above was also based on a simple SEIR model. The SEIR model, one of the basic and commonly used deterministic models of directly transmitted infectious diseases, assumes that births, deaths, and other demographic variables, such as the level of human mobility, vaccine coverage, and the age-structure effect, are mainly negligible. On the other hand, for relatively small groups, such as schools and sports teams, the assumption of a homogeneous population is more appropriate, and our model can be safely applied to decision-making in such small groups. In addition, compartmental models, such as the SEIR model, have the advantage that the test’s sensitivity can be set according to the infection status, to assess the effectiveness of testing. In fact, several previous studies used SEIR models to discuss and evaluate the effectiveness of testing in small groups [12, 11, 23, 22]. However, the present study may function as a preliminary stage in studies based on advanced models, which may help to understand overall trends before conducting more detailed analyses using such models [9]. Furthermore, a number of studies have applied simple SEIR models to discuss the transmission of infectious diseases in large populations [6, 18]; and even if a given population should have a heterogeneous structure, if the transition of the infection status can be explicitly described, the dynamics can be described by the analytical method used in the present study.

Finally, to derive the study’s infection probabilities, we applied the parameters reported by Kucirka *et al*. [14] for the sensitivity and specificity of the two tests. As our results depended on these parameters, we conducted a sensitivity analysis for the parameters. The parameters *a*_1_, *a*_2_, and *a*_*s*_ were randomly sampled from the intervals [0.25, 0.45], [0.50, 0.70], and [0.70, 0.90], respectively. Note that these intervals contain the original values, *a*_1_ = 0.33, *a*_2_ = 0.61, and *a*_*s*_ = 0.80, respectively, and *a*_1_ ≤ *a*_2_ ≤ *a*_*s*_ always holds. Then, the probability of infection on day 0, the respective ratios of the probability of infection on days 5 and 10 to day 0, and the difference between the ratios on days 5 and 10, were calculated. This procedure was repeated 100 times. Table 10 shows the results. On average, for Case (i), the infection rate decreases 17% in the first 5 days, but only 9.8% in the following 5 days. In other words, the infection rate drops dramatically in the first 5 days, but not much afterward. This pattern is also observed for the maximum and minimum infection rates, and for Case (ii); and it coincides with the results discussed in the ‘Reduction of the isolation period’ section of Discussion. In summary, though there are some differences in the resulting values, depending on the sensitivity setting of the tests, it can be confirmed that the pattern of the results does not change.

**Table 10:** Results of sensitivity analysis for the sensitivity of the two tests. Day 0 stands for the probability of infection on day 0. Day 5*/*Day 0 and Day 10*/*Day 0 are the ratios of the probability of infection on days 5 and 10 to day 0, respectively, and the last column shows their difference. Average, Min., and Max. indicate the results when the average, minimum, and maximum values of the 100-repetition simulation are applied.

|  |  | Day 0 | Day 5/Day 0 | Day 10/Day0 | The 2nd column – the 3rd column |
| --- | --- | --- | --- | --- | --- |
| Case(i) | Average | 0.0090 | 0.17 | 0.077 | 0.098 |
|  | Min. | 0.0079 | 0.098 | 0.043 | 0.053 |
|  | Max. | 0.0010 | 0.25 | 0.11 | 0.14 |
| Case(ii) | Average | 0.0082 | 0.16 | 0.080 | 0.078 |
|  | Min. | 0.00070 | 0.092 | 0.049 | 0.043 |
|  | Max. | 0.0093 | 0.22 | 0.11 | 0.11 |

## Conclusions

The present study investigated, in the context of COVID-19 transmission:

- the relationship between the length of the isolation period and the expected infection probability; and
- the expected infection probability with negative antigen test certification.

With regard to the first item, the study results suggest that a five-day isolation period is effective from the view of risk management; and that, if a longer period is contemplated, then both the effect on the risk reduction and the possible negative effects due to the longer isolation should be considered. Regarding the second item, the results provide quantitative information that may be useful for the decision-making of officials in managerial positions.

## Data Availability

All data produced in the present study are available upon reasonable request to the authors.

https://github.com/junbow52/infection_probability

## Author contributions statement

JT made substantial contributions to the design of the study and the analysis, and drafted the article. MM made substantial contributions to the conception and design of the study, and substantively revised the draft. MK made substantial contributions to the design of the study and the analysis. WN, TY, and SI made substantial contributions to the conception and design of the study. All the authors have approved the submitted version and agreed both to be personally accountable for the author’s own contributions, and to ensure that questions related to the accuracy or integrity of any part of the study, even ones in which the author was not personally involved, are appropriately investigated and resolved, with the resolution documented in the literature.

## Additional information

Among the outside support for the study, WN and TY report a relationship with Kao Corporation (funding grants); and a relationship with the Nippon Professional Baseball Organization, Yomiuri Giants, Tokyo Yakult Swallows, Japan Professional Football League, and Japan Professional Basketball League (funding grants).

Other activities: the study was conducted as part of a comprehensive research project, comprising members from two private companies, Kao Corporation and NVIDIA Corporation, Japan; however, no authors in the study belong to these companies. MM, MK, WN, TY, and SI have attended the new Coronavirus Countermeasures Liaison Council, jointly established by the Nippon Professional Baseball Organization and Japan Professional Football League, as experts without remuneration. WN and TY are advisors to the Japan National Stadium and Japan Professional Football League. The other authors declare no competing interests. The findings and conclusions of this article are solely the responsibility of the authors, and do not represent the official views of any institution.

## Data availability

A Mathematica notebook for calculating infection probabilities are available at https://github.com/junbow52/infection_risk/.

## References

[1] B. Adamson, R. Sikka, A. L. Wyllie, and P. Premsrirut. Discordant SARS-CoV-2 PCR and Rapid Antigen Test Results When Infectious: A December 2021 Occupational Case Series. medRxiv, 2022. doi:10.1101/2022.01.04.22268770.

[2] R. M. Anderson and R. M. May. Infectious Diseases of Humans: Dynamics and Control. Oxford Science Publications. Oxford University Press, Oxford; New York, 1991. ISBN 978-0-19-854599-6.

[3] T. K. Burki. Lifting of COVID-19 restrictions in the UK and the Delta variant. The Lancet Respiratory Medicine, 9(8):e85, Aug. 2021. ISSN 22132600. doi:10.1016/S2213-2600(21)00328-3.

[4] E. Callaway and H. Ledford. How bad is Omicron? What scientists know so far. Nature, 600(7888):197–199, Dec. 2021. ISSN 0028-0836, 1476-4687. doi:10.1038/d41586-021-03614-z.

[5] O. Diekmann, J. Heesterbeek, and J. Metz. On the definition and the computation of the basic reproduction ratio R 0 in models for infectious diseases in heterogeneous populations. Journal of Mathematical Biology, 28(4): 365–382, 1990. ISSN 0303-6812, 1432-1416. doi:10.1007/BF00178324.

[6] Z. Du, A. Pandey, Y. Bai, M. C. Fitzpatrick, M. Chinazzi, A. Pastore y Piontti, M. Lachmann, A. Vespignani, B. J. Cowling, A. P. Galvani, and L. A. Meyers. Comparative cost-effectiveness of SARS-CoV-2 testing strategies in the USA: A modelling study. The Lancet Public Health, 6(3):e184–e191, 2021. ISSN 24682667. doi:10.1016/S2468-2667(21)00002-5.

[7] W. He, G. Y. Yi, and Y. Zhu. Estimation of the basic reproduction number, average incubation time, asymptomatic infection rate, and case fatality rate for COVID-19: Meta-analysis and sensitivity analysis. Journal of Medical Virology, 92(11):2543–2550, Nov. 2020. ISSN 0146-6615, 1096-9071. doi:10.1002/jmv.26041.

[8] X. He, E. H. Y. Lau, P. Wu, X. Deng, J. Wang, X. Hao, Y. C. Lau, J. Y. Wong, Y. Guan, X. Tan, X. Mo, Y. Chen, B. Liao, W. Chen, F. Hu, Q. Zhang, M. Zhong, Y. Wu, L. Zhao, F. Zhang, B. J. Cowling, F. Li, and G. M. Leung. Temporal dynamics in viral shedding and transmissibility of COVID-19. Nature Medicine, 26(5):672–675, May 2020. ISSN 1078-8956, 1546-170X. doi:10.1038/s41591-020-0869-5.

[9] H. W. Hethcote. The mathematics of infectious diseases. SIAM Review, 42(4):599–653, Jan. 2000. ISSN 0036-1445, 1095-7200. doi:10.1137/S0036144500371907.

[10] L. Jansen, B. Tegomoh, K. Lange, K. Showalter, J. Figliomeni, B. Abdalhamid, P. C. Iwen, J. Fauver, B. Buss, and M. Donahue. Investigation of a SARS-CoV-2 B.1.1.529 (Omicron) Variant Cluster — Nebraska, Novem-ber–December 2021. MMWR. Morbidity and Mortality Weekly Report, 70(5152):1782–1784, Dec. 2021. ISSN 0149-2195, 1545-861X. doi:10.15585/mmwr.mm705152e3.

[11] M. Kamo, M. Murakami, and S. Imoto. Effects of test timing and isolation length to reduce the risk of COVID-19 infection associated with airplane travel, as determined by infectious disease dynamics modeling. Microbial Risk Analysis, 20:100199, 2022. ISSN 23523522. doi:10.1016/j.mran.2021.100199.

[12] M. Kamo, M. Murakami, W. Naito, J.-i. Takeshita, T. Yasutaka, and S. Imoto. COVID-19 testing systems and their effectiveness in small, semi-isolated groups for sports events. PLOS ONE, 17(3):e0266197, 2022. ISSN 1932-6203. doi:10.1371/journal.pone.0266197.

[13] W. O. Kermack and A. G. McKendrick. A contribution to the mathematical theory of epidemics. Proceedings of the Royal Society of London. Series A, 115(772):700–721, 1927. ISSN 0950-1207, 2053-9150. doi:10.1098/rspa.1927.0118.

[14] L. M. Kucirka, S. A. Lauer, O. Laeyendecker, D. Boon, and J. Lessler. Variation in False-Negative Rate of Reverse Transcriptase Polymerase Chain Reaction–Based SARS-CoV-2 Tests by Time Since Exposure. Annals of Internal Medicine, 173(4):262–267, Aug. 2020. ISSN 0003-4819, 1539-3704. doi:10.7326/M20-1495.

[15] T. Kuniya. Evaluation of the effect of the state of emergency for the first wave of COVID-19 in Japan. Infectious Disease Modelling, 5:580–587, 2020. ISSN 24680427. doi:10.1016/j.idm.2020.08.004.

[16] M. J. Mina, R. Parker, and D. B. Larremore. Rethinking Covid-19 test sensitivity — A strategy for containment. New England Journal of Medicine, 383(22):e120, Nov. 2020. ISSN 0028-4793, 1533-4406. doi:10.1056/NEJMp2025631.

[17] Ministry of Health, Labour and Welfare. Q&A on the new coronavirus (for the general public) (In Japanese). https://www.mhlw.go.jp/stf/seisakunitsuite/bunya/kenkou_iryou/dengue_fever_qa_00001.html#Q3-3.

[18] M. Z. Ndii and Y. A. Adi. Modelling the transmission dynamics of COVID-19 under limited resources. Communications in Mathematical Biology and Neuroscience, 62:1–24, 2020. ISSN 20522541. doi:10.28919/cmbn/4912.

[19] J. L. Prince-Guerra, O. Almendares, L. D. Nolen, J. K. L. Gunn, A. P. Dale, S. A. Buono, M. Deutsch-Feldman, S. Suppiah, L. Hao, Y. Zeng, V. A. Stevens, K. Knipe, J. Pompey, C. Atherstone, D. P. Bui, T. Powell, A. Tamin, J. L. Harcourt, P. L. Shewmaker, M. Medrzycki, P. Wong, S. Jain, A. Tejada-Strop, S. Rogers, B. Emery, H. Wang, M. Petway, C. Bohannon, J. M. Folster, A. MacNeil, R. Salerno, W. Kuhnert-Tallman, J. E. Tate, N. J. Thornburg, H. L. Kirking, K. Sheiban, J. Kudrna, T. Cullen, K. K. Komatsu, J. M. Villanueva, D. A. Rose, J. C. Neatherlin, M. Anderson, P. A. Rota, M. A. Honein, and W. A. Bower. Evaluation of Abbott BinaxNOW Rapid Antigen Test for SARS-CoV-2 Infection at Two Community-Based Testing Sites — Pima County, Arizona, November 3–17, 2020. MMWR. Morbidity and Mortality Weekly Report, 70(3):100–105, Jan. 2021. ISSN 0149-2195, 1545-861X. doi:10.15585/mmwr.mm7003e3.

[20] D. Sleat, K. Innes, and I. Parker. Are vaccine passports and covid passes a valid alternative to lockdown? BMJ, page 2571, Nov. 2021. ISSN 1756-1833. doi:10.1136/bmj.n2571.

[21] The Japanese professional soccer league. J.LEAGUE PUB Report 2021 (in Japanese). https://jlib.j-league.or.jp/-site_media/media/content/70/1/html5.html#page=1.

[22] X. Wang, Y. Cai, B. Zhang, X. Zhang, L. Wang, X. Yan, M. Zhao, Y. Zhang, and Z. Jia. Cost-effectiveness analysis on COVID-19 surveillance strategy of large-scale sports competition. Infectious Diseases of Poverty, 11 (1):32, 2022. ISSN 2049-9957. doi:10.1186/s40249-022-00955-3.

[23] W. Zhu, J. Feng, C. Li, H. Wang, Y. Zhong, L. Zhou, X. Zhang, and T. Zhang. COVID-19 risk assessment for the Tokyo Olympic Games. Frontiers in Public Health, 9:730611, 2021. ISSN 2296-2565. doi:10.3389/fpubh.2021.730611.

